# COVID-19 genomic susceptibility: Definition of *ACE2* variants relevant to human infection with SARS-CoV-2 in the context of ACMG/AMP Guidance

**DOI:** 10.1101/2020.05.12.20098160

**Authors:** Claire L Shovlin, Marcela P. Vizcaychipi

## Abstract

**Background:** Mortality remains very high and unpredictable in CoViD-19, with intense public protection strategies tailored to preceived risk. Males are at greater risk of severe CoViD-19 complications. Genomic studies are in process to identify differences in host susceptibility to SARS-CoV-2 infection.

**Methods:** Genomic structures were examined for the *ACE2* gene that encodes angiotensin-converting enzyme 2, the obligate receptor for SARS-CoV-2. Variants in 213,158 exomes/genomes were integrated with ACE2 protein functional domains, and pathogenicity criteria from the American Society of Human Genetics and Genomics/Association for Molecular Pathology.

**Results:** 483 variants were identified in the 19 exons of *ACE2* on the X chromosome. All variants were rare, including nine loss-of-function (potentially SARS-CoV-2 protective) alleles present only in female heterozygotes. Unopposed variant alleles were more common in males (262/3596 [7.3%] nucleotides) than females (9/3596 [0.25%] nucleotides, *p*<0.0001). 37 missense variants substituted amino acids in SARS-CoV-2 interacting regions or critical domains for transmembrane ACE2 expression. Four upstream open reading frames with 31 associated variants were identified. Excepting loss-of-function alleles, variants would not meet minimum criteria for classification as ‘Likely Pathogenic/beneficial’ if differential frequencies emerged in patients with CoViD-19.

**Conclusions:** Males are more exposed to consequences from a single variant *ACE2* allele. Common risk/beneficial alleles are unlikely in regions subject to evolutionary constraint. *ACE2* upstream open reading frames may have implications for aminoglycoside use in SARS-CoV-2-infected patients. For this SARS-CoV-2-interacting protein with pre-identified functional domains, pre-emptive functional and computational studies are encouraged to accelerate interpretations of genomic variation for personalised and public health use.

## INTRODUCTION

The human population is currently undergoing natural selection for genomic variants that were not deleterious prior to the SARS-CoV-2 pandemic. COVID-19 infection can result in mild sequelae, but life-threatening disease phenotypes are common [1–7]. Despite intensive public health measures, by June 2020 the disease had claimed more than 380,000 lives [8].

The spectrum of COVID-19 disease severity strongly points towards differences in host genetic susceptibility, and genomic studies are pending [9]. An understanding of who may be more, or less, at risk from SARS-CoV-2 is important for public and personalised health strategies. The most obvious initial questions regard males, with Italian [4] and UK [5] intensive care units reporting striking male excesses in >10,000 admissions, and males displaying more severe biomarkers at all ages [6].

SARS-CoV-2 has an obligate host receptor, transmembrane angiotensin-converting enzyme 2 (ACE2) [1], providing opportunities to examine potential genetic risk. ACE2 is a single membrane-spanning protein with a complex, hinge-bending extracellular structure [10], and normally catalyses the degradation of angiotensin II [11]. Angiotensin II degradation is a crucial regulatory step which has multiple and essential cardio-protective consequences through modification of the renin-angiotensin-aldosterone system (RAAS) [11]. Recently, the crystal structure of SARS-CoV-2 virus interacting with ACE2 was solved to 2.5-Å resolution [12]. The viral Spike (S) envelope protein embeds into the surface of the human host cell, and is cleaved to subunits S1 and S2 (‘primed’) by TMPRSS2, augmented by cathepsin B/ L [13]. This process enables S1 to participate in receptor recognition, and S2 in membrane fusion. As detailed by Wang and colleagues, the S1 C-terminal domain (CTD) interacts with a single membrane-bound molecule of host angiotensin-converting enzyme 2 (ACE2), resulting in internalisation of the complex by the cell [12]. The viral-receptor engagement was shown to be dominated by a series of strong polar contacts formed between specific viral and ACE2 amino acids, resulting in a solid network of hydrogen-bond and salt bridge interactions at the interface [12].

With these insights, it becomes feasible to begin to address differing host genotypic susceptibility to SARS-CoV-2. The study goals were to define human genetic variation in *ACE2*, and consider in the context of the rigorous, structured approaches recommended by the American College of Medical Genetics and Genomics and the Association for Molecular Pathology (ACMG/AMP) [14].

## METHODS

### ACE2 Reference Sequence

The reference sequence for the human 3596 nucleotide transcript encoding ACE2 (NM_021804, 19-Apr-2020 update), was downloaded from RefSeq [15] and cross mapped to the National Center for Biotechnology Information Genome Reference Consortium Human Builds GRCh37/hg19 [16] and GRCh38/hg38 [17], utilising University of Santa Cruz (UCSC) Genome Browser resources [18,19]. Exon 1 and 2 sequences 5’ to the methionine start codon were translated using ExPASY Translate [20].

### Definition of ACE2 nucleotides relevant to SARS-CoV-2 infection

Amino acids participating in hydrogen bonding with SARS-CoV-2 [13], and immediately adjacent residues, were mapped to the primary amino acid and nucleotide sequences of NM_021804. These, and the NM_021804 annotations for nucleotides encoding the transmembrane domain and protease cleavage sites of ACE2 (limited circulating enzyme is released following cleavage of transmembrane ACE2), were mapped to the two human genome builds in current use, GRCh37/hg19 and GRCh38/hg38.

### Assessment of ACE2 human variation

Variants in all *ACE2* exons and flanking intronic regions identified by versions 2 and 3 of the Genome Aggregation Consortium (gnomAD) were downloaded. These 213,158 exomes and genomes from largely non-overlapping 111,454 male and 101,704 female datasets [21], mapped to GRCh37/hg19 [16] (version 2, 141,456 samples), and GRCh37/hg19 [17] (version 3, 71,702 genomes) [21]. Denoted *ACE2* variants were supplemented using genomic coordinates to retrieve additional variants in the untranslated nucleotides of exons 1 and 19. All variants were used to align human genome builds GRCh37/hg19 and GRCh38/hg38 across the *ACE2* locus.

### ACE2 Variant Interpretation

The ACMG/AMP Standards and Guidelines [14] describe a structured approach to DNA variants, integrating evidence in favour or against a functional consequence/pathogenicity. Multiple separate criteria are evaluated for a particular gene variant based on allele frequency, molecular/protein functional predictions, patient phenotypic information, segregation studies in families, and other data from reputable sources [14]. For Pathogenicity (P) there are categories of very strong [PVS]; strong [PS]; moderate [PM] and supporting [PP] criteria of evidence. Separate categories of evidence support Benign (B) status, with standalone [BA], strong [BS] and supporting [BP] criteria for the absence of impact on the function or phenotype under study [14]. Criteria met by the variant are evaluated against rules defining minimum requirements to meet ‘Pathogenic’, ‘Likely Pathogenic’, ‘Likely Benign’ or ‘Benign’ status. Any variant not meeting or exceeding the minimum requirements for a ‘Likely’ designation is classified as a variant of uncertain significance (VUS), and considered non-actionable for clinical care [14].

For COVID 19, variant alleles increasing expression of transmembrane ACE2 protein, or otherwise enhancing capacity to internalise SARS-Co-V into the host cell, may enhance risk. Conversely, loss-of-function variants predict lower COVID 19 risk. Variants with no functional impact would be considered as a VUS or Likely Benign with respect to SARS-CoV-2 infection. For the current study, variants were defined as meeting the ACMG/AMP criterion PVS1 for pathogenic/functional impact (in terms of a COVID resistant state) if they clearly resulted in loss-of-function (null) alleles [14]. Missense variants resulting in amino acid substitutions were evaluated particularly with respect to SARS-CoV-2 receptor status [12, 22].

### Data Analysis

Aligned data were uploaded to STATA IC 15.1 (Statacorp, Texas, US) in order to generate descriptive statistics, perform two way comparisons using χ^2^ and Mann Whitney tests, and generate graphs.

## RESULTS

### ACE2 structure

The human *ACE2* gene comprises 19 exons which span 41.9kb of the X chromosome (*Figure 1*). The 24 key amino acids for viral-receptor engagement [12] are encoded by nucleotides in *ACE2* exons 2, 3, 9 and 10. The stalk cleavage site recognised by ADAM-17, a disintegrin and metalloproteinase domain protein, is encoded by exon 16. The stalk cleavage site recognised by transmembrane protease, serine 2 TMPRSS2 (which also cleaves the viral Spike protein to subunits S1 and S2 [13]) and by transmembrane protease, serine 11D, is encoded by exons 17 and 18 (*Figure 1*).

**Figure 1:**
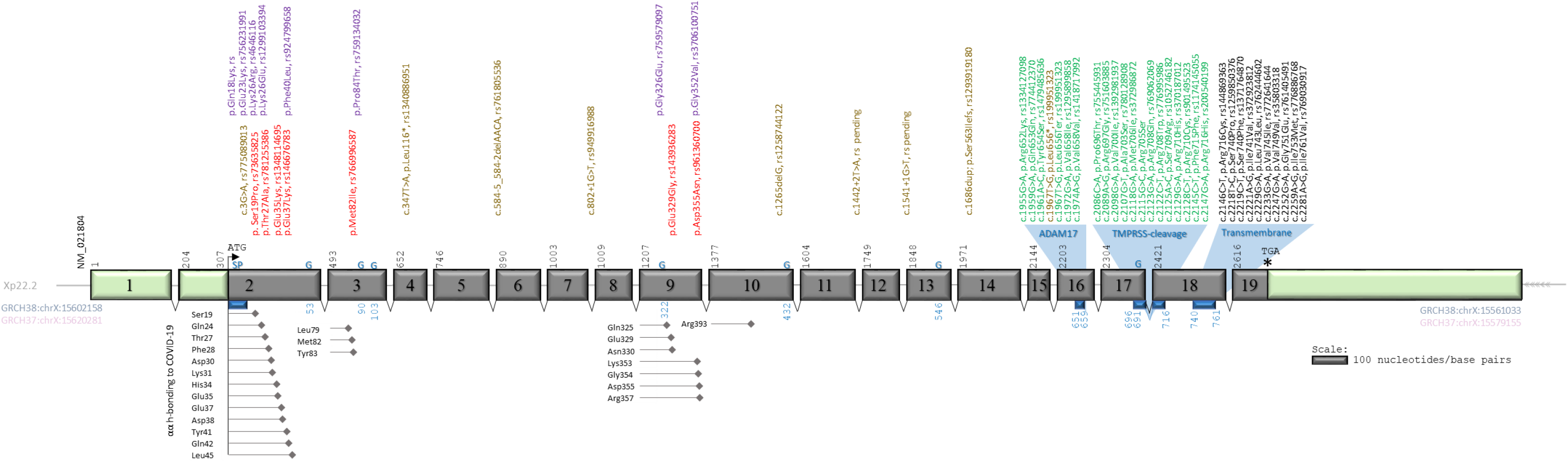
*ACE2* and key variants. The 19 exons of the main transcript of *ACE2* (NM_021804), sited on the X chromosome, and drawn as boxes to scale, with stylised untranslated regions (pale green), intervening introns and splicing events. Black numbers indicate exon first nucleotides, with the ATG start codon and TGA natural stop codon highlighted. Blue text indicates nucleotides encoding the signal peptide (SP), asparagines that are N-glycosylated (G), the transmembrane domain (TM), and two essential regions for cleavage by ADAM-17, and by transmembrane proteases, serine 2 (TMPRSS2) and serine 11D (TMPRSS11D). Below the exons, amino acids (αα) numbered in blue refer to host ACE2 domains, with amino acids that hydrogen (H) -bond with the SARS-CoV-2 virus Spike protein CTD listed vertically, with precise position indicated at the point of the adjacent diamond. Above the exons, variants most likely to change COVID-19 susceptibility are described by coding nucleotide number (c. from methionine start i.e. +307 compared to exon 1 nucleotide 1), consequence (p. protein sequence; *, stop gain; fs, frameshift), and rs designation if available. Text is coloured as follows: Brown, loss of function alleles (start loss, stop gain, frameshift, and splice consensus variants); red, variants replacing amino acid residues directly hydrogen-bonding with SARS-CoV-2; purple, variants replacing an amino acid adjacent to a SARS-CoV-2 hydrogen bonding amino acid; green, variants affecting the cleavage sites, and black, variants affecting the transmembrane domain. Variants are also presented in *Table 1*.

### Coding DNA variants in ACE2

3596 transcribed nucleotides are spliced to form the ACE2 messenger (m)RNA. In these and flanking splice site nucleotides, 483 variants were identified in gnomAD [21], with 305 (63.1%) present in only one of the two datasets. The variants were distributed across the *ACE2* gene, and as shown in *Figure 2*, more than 90% were very rare, affecting fewer than 0.01% of the population. The twelve most common variants are annotated in *Figure 2*. Six of these resulted in amino acid substitutions, four were synonymous variants where the nucleotide sequence still encoded the original amino acid, and two were in the 3’ untranslated region of exon 19.

**Figure 2:**
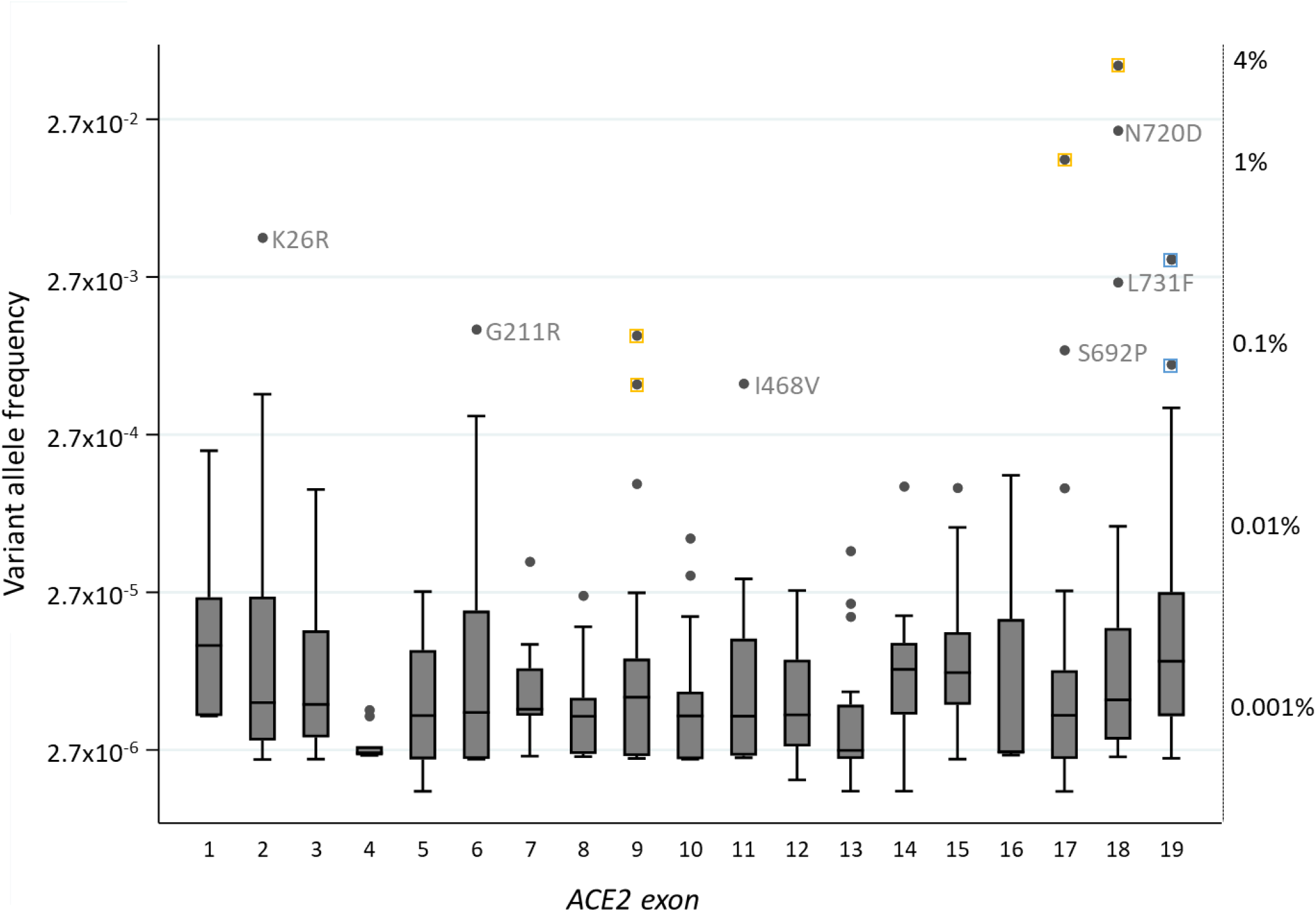
Allele frequencies of *ACE2* exonic variants. The allele frequency of all variants, grouped by exon and plotted on a logarithmic scale. Individual mean allele frequencies were calculated for variants present in more than one database, before calculation of median frequency of all variant alleles in each exon. Corresponding percentage frequencies are indicated on the right, anchored by key individual and median allele frequencies. Box plots indicate median number and interquartile range, error bars 95^th^ percentiles, and dots are outliers. For the most common variants, yellow squares indicate synonymous variants where the nucleotide changes would not result in amino acid substitutions, and blue, non-coding variants in the 3’ untranslated region in exon 19. On the y axis, 2.7 represents e, the base of the natural logarithm that approximates to 2.71828.

### Males are more likely to have an unopposed variant of ACE2

The implications of variant alleles differ for males with their single X chromosome, and single copy of *ACE2*, compared to females with two. Across the 19 exons of *ACE2*, only 9/3596 (0.25%) nucleotides were homozygous variant in at least one female (*Figure 3A*) compared to 262/3596 (7.3%) present in a hemizygous state in at least one male with their single X-chromosome (*Figure 3B*, χ ^2^*p*<0.0001).

**Figure 3:**
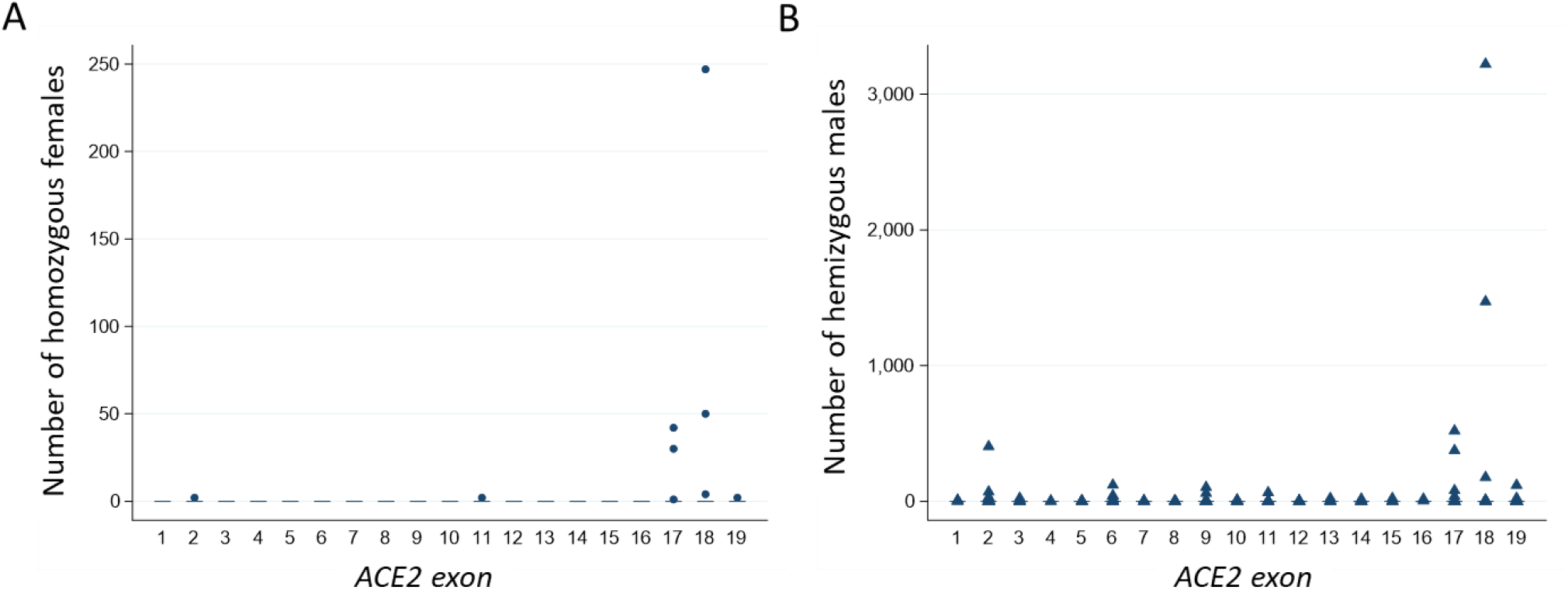
Number of individuals without a normal *ACE2* sequence. **A)** The number of females homozygous for (i.e. having two copies of) an *ACE2* variant, plotted by the exon in which the nucleotide(s) are sited. **B)** The number of males hemizygous for *ACE2* variants (i.e. having their only copy of the *ACE2* sequence a variant) plotted by the exon in which the relevant nucleotide(s) are sited. Note the difference in y-axis scales between **A)** and **B)**, similar patterns of distribution (reflecting overall variant allele frequency in the population, data not shown), and that most variation occurs in exons 17 and 18.

### ACE2 exhibits few coding variants that generate an ACE2 loss-of-function allele

For ACE2, gnomAD version 2 and 3 databases included one whole gene duplication. There were 9 clear loss-of-function variant alleles that would prevent any production of ACE2 protein/SARS-CoV-2 receptor. No full or multi-exon deletions were identified, but several frameshift insertions/deletions resulted in transcript ablation, and other loss-of-function variants resulted from start codon loss, stop codon gains, or disruption of ±2 splice site consensus sequences (*Table 1, Figure 1*).

**Table 1:**
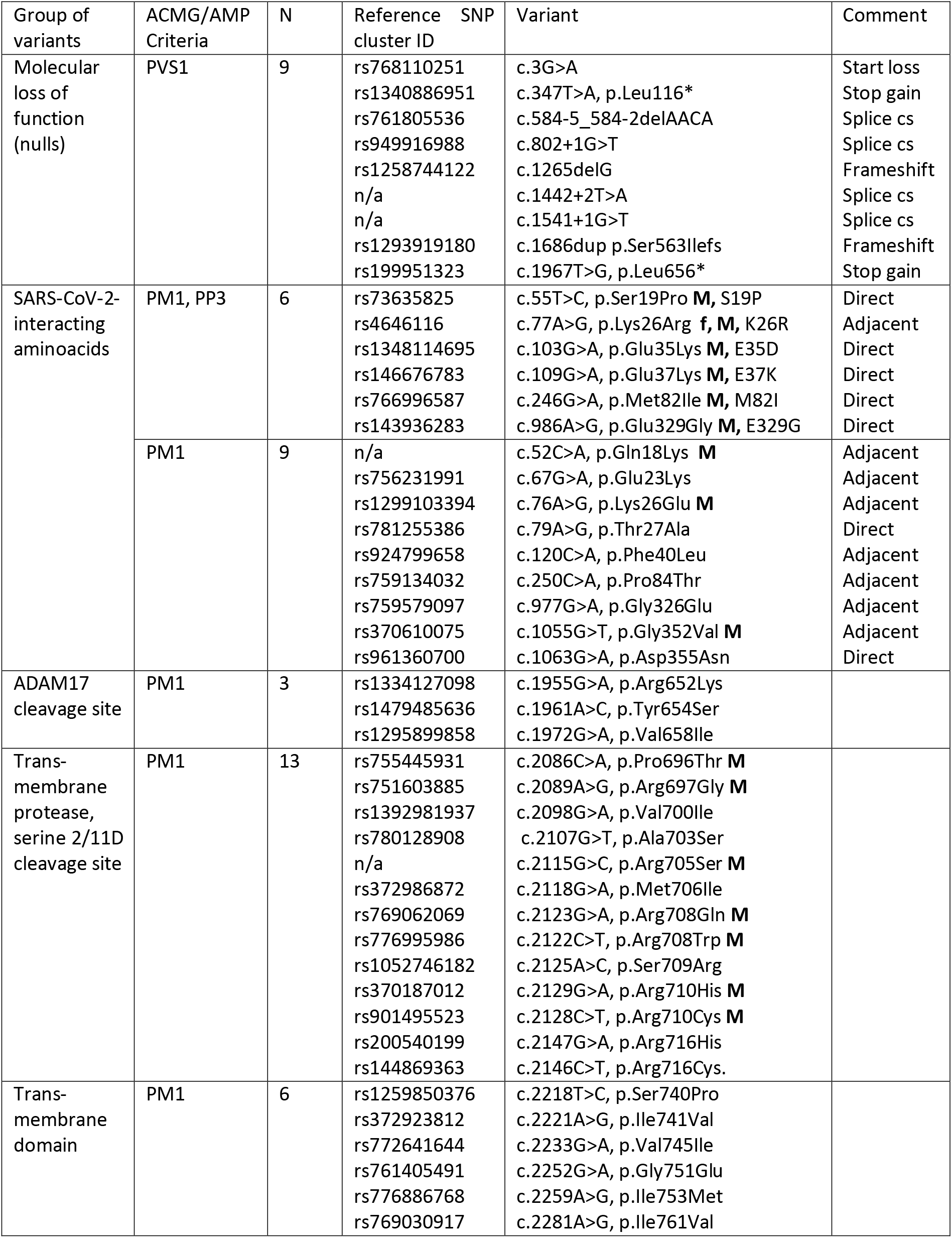
List of variants in key domains of ACE2. Details of the variants falling into particular categories as defined by meeting particular criteria towards pathogenicity. N, number of variants in category; cs, consensus; n/a, not available. ACMG/AMP, American College of Medical Genetics and Genomics and the Association for Molecular Pathology very strong (PVS), moderate (PM) and supporting (PP) evidence for pathogenicity. All variants were present in the heterozygous state in females. **f**, present in a homozygous female. **M**, present in a hemizygous male. 1 letter code variant descriptors also provided if mentioned in text or *Figure 2*.

The loss-of-function variants were only found in the heterozygous state in females who had a second, normal (“wild-type”) allele on their other X chromosome.

### ACE2 exhibits few coding variants substituting amino acids that hydrogen bond with SARS-CoV-2

The ACE2 peptide sequence comprises 805 amino acids, 24 of which participate in hydrogen bonding with SARS-CoV-2 [12]. The gnomAD databases indicated that 7 of the 24 SARS-Co-V-interacting amino acids are substituted by other amino acids in members of the human race, and a further 8 substitutions affect the immediate flanking amino acids (*Table 1, Figure 1*). The individual frequencies of these variant alleles ranged from fewer than 1 in 100,000 to 0.4% (1 in 250) of the gnomAD populations. The most common variant, *ACE2* c.77A>G, p.(Lys26Arg)/K26R, substitutes the residue immediately preceding the SARS-CoV-2-bonding Thr27 and Phe28.

Across the 213,158 datasets in gnomAD versions 2 and 3, two females (0.002%) were homozygous for c.77A>G, p.(Lys26Arg)/K26R, and 429 males (0.39%) had a single unopposed copy of one of eight variants listed in *Table 1*.

### ACE2 exhibits few variants substituting amino acids involved in maintenance of the transmembrane state

Thirty-one amino acids are in the ACE2 stalk cleavage site or transmembrane domain (*Figure 1*). Three missense variants were identified in nucleotides that encode the ADAM17 cleavage site (*Table 1, Figure 1*), all with population allele frequencies < 0.0046%. Thirteen missense variants were identified in nucleotides that encode the TMPR cleavage site (*Table 1, Figure 1*). For these, respective allele frequencies were < 0.026%. Similarly for the transmembrane domain, the 6 identified missense variants (*Table 1, Figure 1*) had a maximal allele frequency <0.011%.

For these 22 variants, in the 213,158 gnomAD datasets, no female was homozygous for any substitution. 58 males (0.05%) had a single unopposed copy of at least one variant (*Table 1*).

### Upstream open reading frames in the ACE2 gene

The regions in *ACE2* exons 1 and 2 that are 5’ to the transcriptional start site are illustrated in *Figure 4*. On ExPASy translation, the first exon and the 5’ region of the second exon contained 4 non-overlapping upstream open reading frames (uORFs). The first uORF is in-frame with the main transcript, the second, third and fourth are in the +1 reading frame. As indicated in *Figure 4*, 12 variants were in the three of the uORFs, a further 7 variants in the uORFs 5’untranslated regions (UTRs), and 14 in the second exon 5’ UTR for the main transcript. All were rare, with individual allele frequencies <0.028%.

**Figure 4:**
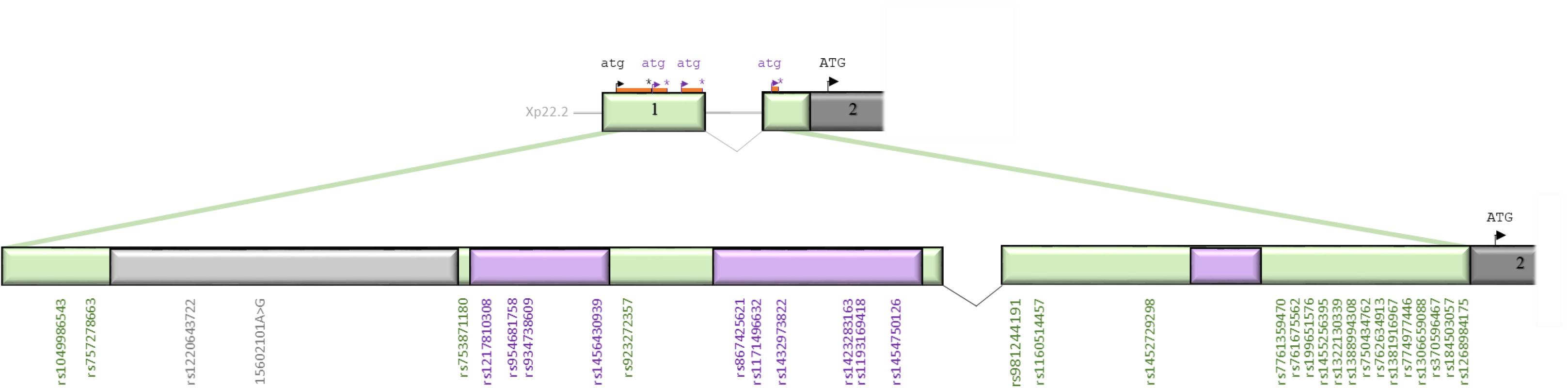
Variants in the 5’ “untranslated” region of *ACE2*. *ACE2* exon 1 and the start of exon 2 with upstream open reading frames (ORFs) to scale, stylised intervening intron/splicing event, and variants approximately positioned. Note that compared to Figure 1, the 5’ “untranslated” region has been deconstructed into open reading frames and 5’/intervening untranslated regions. Grey indicates the in-frame ORF, with variants listed in grey text. Lilac indicates alternate reading frame ORFs with variants listed in purple text. Pale green indicates the intervening untranslated regions, with the relevant variants listed in green text.

No variant was present in a homozygous state in females. In total, 46 males (0.041%) were hemizgous for a variant in exon 1 (rs1049986543, rs954681758, rs934738609, rs1432973822, rs867425621), or in the untranslated regions of exon 2 (rs981244191, rs1452729298, rs370596467, rs762634913, rs1388994308, rs1455256395).

### ACMG/AHP Classification Considerations

The 9 loss-of-function alleles met the ACMG PVS1 criterion in support of “pathogenicity”, noting this would be for the beneficial state of resistance to SARS-CoV-2 infection, since each would prevent any ACE2/SARS-CoV-2 receptor production. The 15 variant alleles for amino acids adjacent and directly interacting with SARS-CoV-2, and 22 variant alleles affecting amino acids in the transmembrane domain, or membrane-proximal cleavage sites, could meet the ACMG/AHP moderate criterion of PM1 in terms of COVID-19, on the basis of affecting a critical domain. In this setting, variants that impaired ACE2/SARS-CoV-2 interaction, prevented ACE2 membrane anchoring, or augmented ACE2 cleavage would be predicted to enhance resistance to SARS-CoV-2 infection, while those that increased ACE2/SARS-CoV-2 interactions, or impeded cleavage of ACE2 would be predicted to enhance susceptibility.

Meeting a single criterion does not meet the minimum requirements for ‘Likely Pathogenic’ [or beneficial] by ACMG/AMP guidance [14]. For the 9 loss-of-function alleles meeting PVS1, at least one further criterion of strong or moderate evidence is required. For the 37 missense variants meeting PM1 due to presence in key functional domains, a ‘Likely Pathogenic’ assignment would require at least two further moderate criteria, or a combination with up to 4 supporting criteria, only one of which can reflect differential allele frequency in disease and non-disease cohorts [14].

## DISCUSSION

We have shown that the *ACE2* gene on the X-chromosome provides a biological rationale why males and females have different risk profiles for COVID-19 infection. Within coding nucleotides, identified variants were rare, with complete loss-of-function (putative SARS-CoV-2-protective) alleles only observed in the heterozygous state in females, and unopposed variants present more commonly in males. The first and second exon upstream open reading frames were identified as a source of ACE2 regulation and variation relevant for patients treated with aminoglycoside antibiotics. Excepting very rare loss-of-function alleles, different patterns of allele frequencies captured in very large COVID-19 sequencing initiatives will be unlikely to meet minimum ACMG/AMP criteria for pathogenicity without further functional data.

The current manuscript utilised gnomAD databases due to their broad population coverage. It is widely considered that variant databases are already saturated for common alleles, although further rare variants are expected to emerge, particularly from previously under-represented populations [23]. Thus, although a limitation of the current study is that not every human genomic database was examined, a wider number of coding *ACE2* variants were retrieved compared to other publications [24], most likely due to combined use of both the GRCh37/hg19-mapping and more recent GRCh37/hg19-mapping gnomAD databases. The study by design was limited to *ACE2*, in order to illustrate principles, but could be expanded to additional potential regions of genomic risk.

Study strengths include highlighting the landscape of genetic variation relevant to SARS-CoV-2, and questioning how strictly putative COVID-19 risk and protective alleles should be defined, particularly if they may become part of public health policies, as for underlying health conditions [25,26]. The pattern from Mendelian disorders was of over-exuberant pathogenicity calls in early research reports, with pathogenicity classifications of many variants later downgraded as assignments became increasingly stringent [14,28]. As a result, high proportion of coding variants in known disease-causing genes are currently clinically non-actionable due to classification as a VUS. This is important because many individuals are “pre-genotyped,” having undergone whole genome sequencing as part of other initiatives [29–31], and will have direct access to open-source research publication of COVID-19 associated variants, before they are subjected to the rigorous oversights applied by Clinical Geneticists.

To facilitate classification if variants are found to be associated with greater or lesser SARS-CoV-2 infection/consequences than expected by chance, *in vitro* modelling of infection rates in ACE2 heterozygous cells would provide evidence towards one of the ACMG/AHP functional studies criteria (PS3). Computational modelling for the small number of specific missense variants at amino acids directly interacting with SARS-Co-V has commenced, and may also meet the ACMG/AMP strong criterion of evidence (PS3): Hussain and colleagues demonstrated that two SARS-Co-V interactions of ACE lysine 353 are predicted to be absent if the apparently ‘distant’ serine 19 is substituted by proline (S19P) [22]. Modelling docking poses of SARS-Co-V-2 suggested intermolecular contacts were also fewer than predicted for E329G, and inter-residual interaction maps indicated that SARS-Co-V interaction with ACE Q42 was absent in E35D, E37K, M82I, and E329G. Additionally, a significant change in the estimated free energy of ACE2 for S19P and K26R implied these may adversely affect protein stability compared to wildtype.[22] Thus PM1 and PP3 (in terms of “pathogenicity” for a COVID-resistant state) would be met for six variants including *ACE2* c.77A>G, p.Lys26Arg, the second most common *ACE2* missense substitution in the human population (*Figure 2*).

Overall, the paucity of loss-of-function alleles in this x-chromosome gene points towards strong evolutionary constraint based on the normal role of ACE2 as a critical homeostatic mediator through the renin angiotensin system [11]. The similarly low allele frequencies for missense substitutions in the SARS-Co-V binding region and stalk cleavage sites suggest these too have been under evolutionary constraint, potentially for the same homeostatic role. In males, the wider consequences of an unopposed loss-of-function allele in this essential transmembrane carboxypeptidase are not yet known, but notably no such males were in the gnomAD datasets. The rare female heterozygotes with a single loss-of-function *ACE2* allele will be protected from total protein absence due to Lyonisation when one female X chromosome is randomly inactivated in each cell [32,33], and may benefit from relative protection from SARS-CoV-2 infection.

Males are therefore more likely to have unopposed risk alleles, but females more likely to benefit from protective alleles better tolerated in the heterozygous state. The rarity of variants in *ACE2* exons indicate that if common resistance or susceptibility alleles to SARS-Co-V emerge, they are unlikely to be found in the *ACE2* coding regions. Currently the Genotype-Tissue Expression (GTEx) project identifies 2,053 expression quantitative trait loci (eQTLs) for *ACE2* based on mapping windows 1Mb up and downstream of the transcription start site [34]. While these loci will likely harbour SARS-CoV-2 genomic susceptibility and resistance alleles, and some are very common, with minor allele frequencies approaching 0.5, the significant linkage disequilibrium implies that selecting individual loci for functional evaluations may be challenging [35].

For immediate clinical considerations, regulation of the *ACE2* uORFs is suggested as a focus for functional assays in the presence and absence of relevant variants. This is because aminoglycoside antibiotics (such as gentamicin, amikacin, neomycin, tobramycin and kanamycin that are used to treat patients including those with COVID-19) are used *in vitro* to manipulate uORFs via ribosomal read-through of termination codons [36]. In general, uORFs are known to be able to stimulate or inhibit translation of main ORFs [37]. *In vitro* studies would provide evidence whether aminoglycosides have no effect, increase or decrease ACE2 transcription and thus whether no effect, beneficial or deleterious effects on disease progression would be predicted.

In conclusion, given the obligate host receptor for SARS-CoV-2 is located on the X chromosome, genetic considerations supported by population-based genotyping further emphasise the importance of increasing targeted and personalised care to males, and highlight priority regions of the human genome for interrogation. These focus not only on *ACE2* amino acids interacting with SARS-CoV-2 and in critical sites that maintain ACE2 in its transmembrane form, but also the upstream reading frames where read-through may be modified by antibiotic treatments used in SARS-CoV-2-infected individuals. Given the large number of COVID-19 host genomics initiatives, differential allele frequencies will probably emerge before variants meet ACMG/AMP criteria for actionable results in patients. Pre-emptive functional studies, and ACMG/AMP-structured approaches to research-based presentation of COVID-19 susceptibility variant results are encouraged.

## Data Availability

Data are available in the primary data sources (references 15-19,21). We are in the process of developing an accessible database that will be open access following peer review publication.

## Acknowledgements

We gratefully acknowledge the research laboratories and bioinformatics groups whose open source materials made these analyses feasible. The work received no specific funding. CLS acknowledges infrastructure support provided by the NIHR Imperial Biomedical Research Centre. The views expressed are those of the authors and not necessarily those of funders, the NHS, the NIHR, or the Department of Health and Social Care.

## Data Availability Statement

Data are available in the primary datasources (references 15-19,21). We are in the process of developing an accessible database that will be open access following peer review publication.

## Ethical Approvals

Ethical approvals were not required as all data reported were already anonymised and in the public domain.

